# LONGITUDINAL CLUSTER ANALYSIS OF HEMODIALYSIS PATIENTS WITH COVID-19 IN THE PRE-VACCINATION ERA

**DOI:** 10.1101/2022.08.20.22279014

**Authors:** Pasquale Esposito, Sara Garbarino, Daniela Fenoglio, Isabella Cama, Leda Cipriani, Cristina Campi, Alessia Parodi, Tiziana Vigo, Diego Franciotta, Tiziana Altosole, Fabrizio Grosjean, Francesca Viazzi, Gilberto Filaci, Michele Piana

**Author notes:** Corresponding Author: Pasquale Esposito, Unit of Nephrology, Dialysis and Transplantation, Department of Internal Medicine, University of Genova and IRCCS Ospedale Policlinico San Martino, Genova, Italy.

## Abstract

Coronavirus disease 2019 (COVID-19) is characterized by a high heterogeneity of clinical presentation and outcomes. This is also true for patients undergoing maintenance hemodialysis (HD), who, due to specific clinical factors and immune status, represent a distinct subgroup of COVID-19 patients.

Starting from this observation in this research letter we tested and validated in two cohorts of HD patients with COVID-19 (derivation and validation cohort, respectively) an innovative model which combines linear mixed effect modeling and cluster analysis on longitudinal.

This study aimed to describe a methodology allowing patient stratification from simple and widely available data.

Our results could be interesting not only to improve COVID-19 management but also to support the application of longitudinal cluster analysis strategy in other clinical settings.

---

The heterogeneity of clinical presentation and outcomes is a common feature of Coronavirus disease 2019 (COVID-19)^1^. This is also true for maintenance hemodialysis (HD) patients, who represent a distinct subgroup of COVID-19 patients^2,3^. Taking advantage of these peculiarities, we investigated the presence of different COVID-19 subtypes in this population using an innovative model which combines linear mixed effect modelling and cluster analysis.

We explored the potentiality of this approach studying patients affected by COVID-19 during the first pandemic waves of 2020, to evaluate the natural history of the infection without the interference of external confounding factors, such as vaccinations and antivirals.

The study was performed according to the Declaration of Helsinki and was approved by the local Ethics Committee (N. Registro CER Liguria: 135/2020). All participants provided written informed consent before enrollment.

The derivation cohort (DC) was constituted by 17 HD patients (66.7±12.3 years 8 males) with diagnosis of COVID-19 during the second pandemic wave of 2020 (October-December). We collected clinical data, general laboratory, and cytokine determinations, and cytofluorimetric analysis of activation and maturation of CD4+ and CD8+ lymphocyte subpopulations (Supplementary Table 1)^4^. Disease presentation severity was scored as 0 (asymptomatic patients), 1 (mildly symptomatic), or 2 (full symptomatic) (see Supplementary Methods). Data were collected at diagnosis and days 7-14-21-28, with each subject having at least two time-points acquisition for each biomarker. Overall, we collected 51 longitudinal observations over 4 time-points on 17 subjects. First, we compared the baseline values of COVID-19 HD, with a sex-age matched control group of 6 HD patients (70.0±9.4 years, 3 males) tested negative for COVID-19 (Supplementary Table 2) and selected the 10 most significant different variables between the two groups and for which at least 90% of COVID-19 HD patients had a baseline acquisition. These variables included: C-reactive protein (CRP), white blood cell, neutrophil and lymphocyte counts, albumin, ferritin serum levels, and IL-1β, IL-8, IL-6, and TNF-α circulating levels.

**Table 1.**
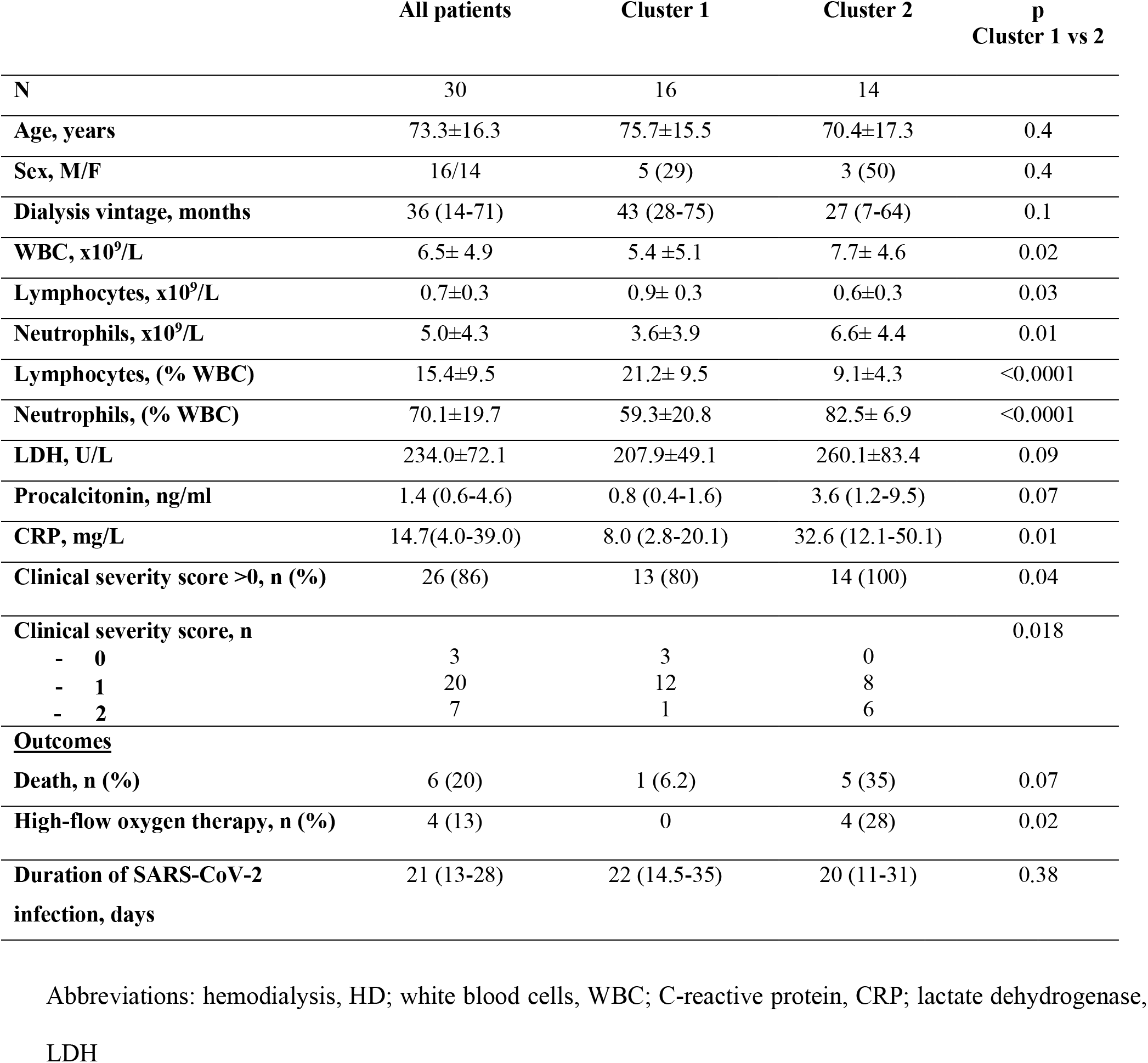
Validation cohort of longitudinal cluster analysis performed in COVID-19 positive HD patients: General characteristic and comparison between patients assigned to different clusters.

Then, we used a linear mixed effect model (LME)^5,6^ on the longitudinal data of all 17 patients of the DC. For each COVID-19 HD patient and each of the 10 features, we obtained a set of two fixed and two random effects (one each for slope and intercept of the LME), which described the biomarker progressions on the overall dataset (fixed effect) and the subject-wise variations (random effects) (Supplementary Figure 1). For both slope and intercept, for each subject, we used the fixed and the random effects as the input of a k-means clustering analysis to identify potential different clusters of COVID-19 HD patients^7^. We set the number of clusters to 3, following a silhouette analysis with Dice distance.

**Figure 1.**
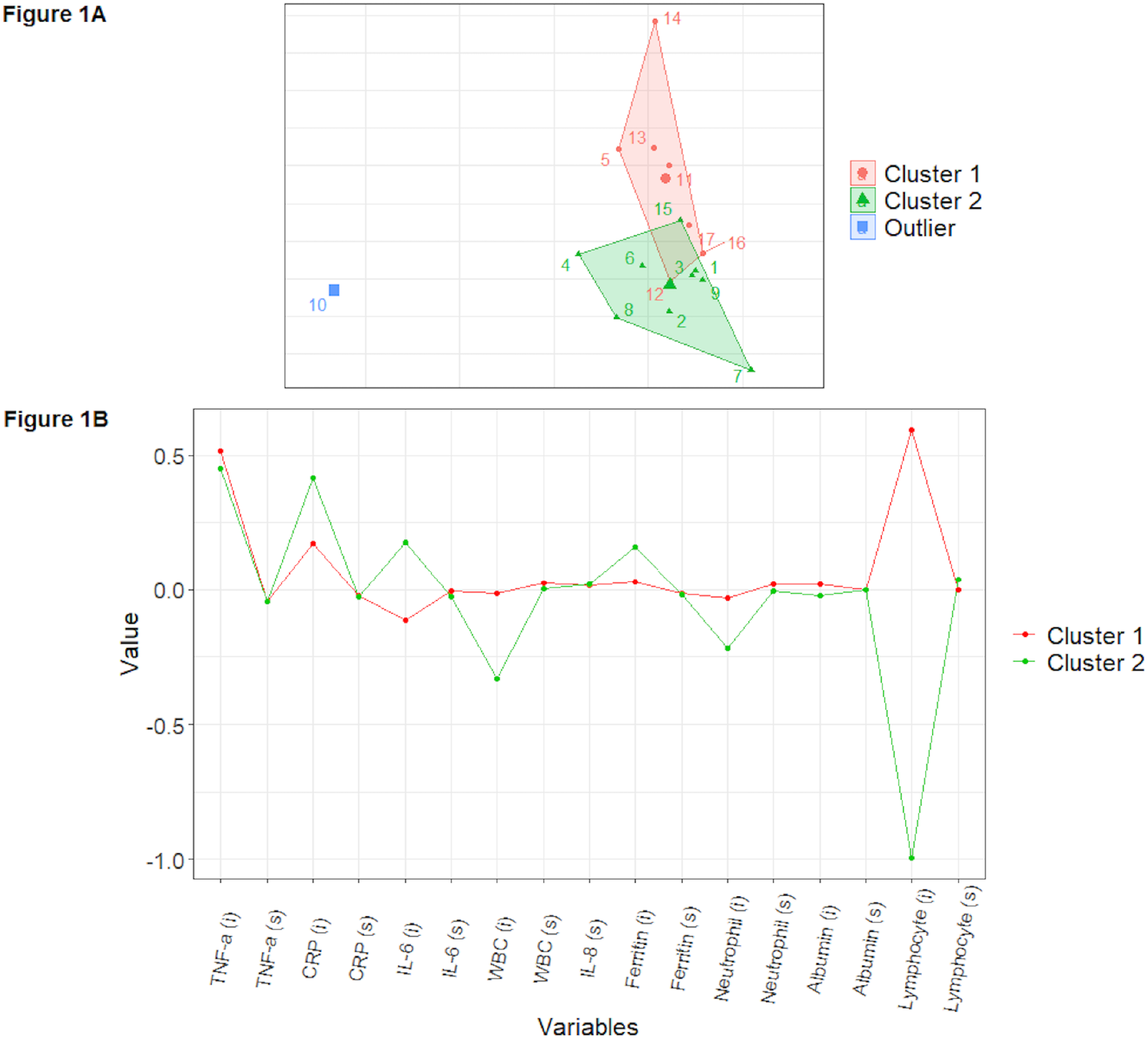
Cluster analysis on longitudinal data performed on the Derivation Cohort. Top panel (1A): result of cluster analysis. Bottom panel (1B): centroid profiles of clusters 1 and 2. Abbreviations: white blood cell count, WBC; C-reactive protein, CRP; interleukin, IL; Tumor necrosis factor-alfa, TNF-*α*; intercept, i; slope, s.

Clustering analysis on the LME parameters returned two well-balanced (7 vs 9 patients) clusters (*cluster 1* and *cluster 2*, Figure 1A) and one cluster consisting of 1 subject, who was excluded from further analysis. The centroid profiles of clusters 1 and 2 (Figure 1B) showed a significant difference in the value of lymphocyte count (p=0.012). The comparison between the two clusters showed that within cluster 1, there were more female patients (p=0.01) presenting significantly higher lymphocyte count (*cluster1* 1.1±0.5 vs *cluster2* 0.4±0.2 ×10^9^/L, p=0.012) and lower lactate dehydrogenase (LDH), CRP (*cluster1* 22.2±16.3 vs *cluster2* 72.6±44.1 mg/dL, p=0.017), and CD8+T memory stem cells (*cluster1* 0.6±0.4 vs *cluster2* 2.3±1.7%, p=0.038), as a possible result of a milder inflammatory and immune response activation (Supplementary Table 3). Then, we tested our clustering strategy in predicting clinical outcomes in a validation cohort (VC), constituted by 30 HD patients (73.3±16.3 years, 16 males) affected by COVID-19 during the first pandemic wave of 2020 (March-April). For each validation subject, we considered the baseline value of 8 “ risk-factor” variables, including sex, age, C-reactive protein, neutrophil, and lymphocyte percentages, procalcitonin, LDH, clinical presentation severity. We used these variables to compute the distances between each validation subject and the cluster centroids to assign each patient to cluster 1 or 2, which were finally constituted by 16 and 14 subjects, respectively, who resulted comparable for general characteristics. We noticed that within cluster 1, disease severity presentation was significantly milder than that observed in cluster 2 (clinical score distribution cluster 1 vs cluster 2, p=0.018). Moreover, in cluster 1, one patient died (6.5%), and none required high-flow oxygen therapy, whereas, in cluster 2, there were five deaths (35%) and four patients (28%) requiring high-flow oxygen supply (p=0.07 and p=0.02, respectively). Finally, there were no significant differences in the duration of the infection (Table 1). In this study, we found that clustering analysis on longitudinal data via linear mixed effect modelling provided information about the differentiation of baseline characteristics and overall disease progression in HD patients with COVID-19 during the first pandemic waves of 2020. This methodology, through the initial analysis of a complex dataset, allowed us to build a final predictive model based on the baseline evaluation of simple clinical and laboratory parameters. Then, although limited by the small number of subjects evaluated and being aware of the rapid evolution of COVID-19, we think our data may support the application of this innovative technique to translate research findings into clinical practice.

## Data Availability

The data underlying this article are available in Harvard Dataverse, at https://doi.org/10.7910/DVN/KDKR4V, (accessed on 16 July 2022).

https://doi.org/10.7910/DVN/KDKR4V

## Supplementary Materials

- Supplementary methods
- Table S1: Full dataset of baseline clinical and laboratory parameters collected from COVID-19 positive and COVID-19 negative HD patients
- Table S2: Comparisons between baseline characteristics of COVID-19 positive and COVID-19 negative HD patients
- Table S3: Baseline characteristics of the derivation cohort of COVID-19 positive HD patients according to cluster assignment
- Table S4: Reagents used for immunofluorescence analyses
- Supplementary Figure 1

