## Supplementary materials for "LONGITUDINAL CLUSTER ANALYSIS OF HEMODIALYSIS PATIENTS WITH COVID-19 IN THE PRE-VACCINATION ERA"

**Supplementary methods**

**Patients**

For this study, we recruited three-weekly maintenance hemodialysis patients with confirmed COVID-19 infection from March 16th to April 30th, 2020 (validation cohort) and from 1st October to 1st December (derivation cohort), enrolled at San Martino, University Hospital of Genoa, Italy. Nasopharyngeal swabs for SARS-CoV-2 were performed on HD patients presenting recent contacts with COVID-19 positive patients and/or with fever or respiratory or gastrointestinal symptoms suspected for COVID-19. The diagnosis was confirmed by positive real-time reverse transcriptase (RT-PCR) assay for SARS-CoV-2.

After the diagnosis, clinical management decisions were left to the attending physicians.

Maintenance HD patients who tested negative for SARS-CoV2 PCR constituted the control group.

**Data collection**

In supplementary Table 1, we report the full set of data collected from each patient at the baseline.

For cytokine determinations, blood samples, collected in polypropylene tubes, were centrifuged at 3,200 rpm for 10 min. Plasma was separated by centrifugation and stored at -20°C until assayed. Then, circulating cytokine levels were determined by Ella automated Immunoassay platform (Protein Simple, USA).

**Immunofluorescence analyses**

Immunofluorescence analyses were performed as previously described (Suppl Ref 1). In particular, 100 µl of peripheral blood was incubated with speciﬁc fluorochrome-conjugated monoclonal antibodies (mAbs) (all purchased from BD Biosciences), as indicated in Supplementary Table 5. The samples were analyzed by a BD Fortessa X20 flow cytometer (BD Biosciences) using the BD FACS Diva™ software version 8.0 (BD Biosciences).

In particular, we considered the following specific phenotypic patterns as markers of T follicular helper (TFH), T helper (Th)1, Th2, Th17, Th1-17, Th9 and Th22 functional T cell subsets, respectively: a) CD4+CXCR5+CD45RA-: TFH; b) CD4+ CXCR3+ CCR4-CCR6- CCR10-: Th1; c) CD4+ CXCR3- CCR4+CCR6-CCR10-: Th2; d) CD4+ CXCR3- CCR4+CCR6+ CCR10- CD161+: Th17; e) CD4+CXCR3+CCR4-CCR6+CD161+: Th17-Th1; f) CD4+CCR4-CCR6+: Th9; g) CD4+ CCR4+CCR6+CCR10+: Th22. Moreover, the sum of the Th17 and Th17-Th1 subsets corresponds to the CD161+CCR6+ population on CD4+ T lymphocytes.

As regards the analysis strategy to follow the maturation of the CD4+ and CD8+T populations, Tcell differentiation has been delineated using a set of canonical markers, i.e. CD45RA, CCR7, CD28, and CD95. Briefly, the differential expression of these markers allows the identification of six subsets in the human peripheral blood: naive (T_N_), stem cell memory (T_SCM_), central memory (T_CM_), transitional memory (T_TM_), effector memory (T_EM_), and terminal effector (T_TE_).

**General statistical methods**

Data are presented as mean ± standard deviation (SD) or interquartile ranges (IQR), if not normally distributed (as evaluated by Shapiro Test). Mann-Whitney test was used to assess the differences among patients affected by COVID-19 and control group, and among patients of different clusters. Proportions for categorical variables were compared using the Fisher’s test. A 2-tailed P value < 0.05 was considered statistically significant.

**Statistical methods for the derivation cohort**

To investigate the presence of different COVID-19 HD subtypes, we used an innovative model which combines linear mixed effect modelling and cluster analysis. The model was implemented in R (https://www.rstudio.com/).

First, we performed a feature reduction procedure by selecting the 10 features whose distribution showed at the same time the highest inter-group difference (COVID-19 HD and HD without COVID-19) and the lowest intra-group variability by means of a Mann-Whitney test (Suppl Ref 2), and for which at least 90% of COVID-19 HD patients had a baseline acquisition. The variables were: C-reactive protein (CRP), white blood cell (WBC), neutrophil and lymphocyte counts, albumin, ferritin serum levels, and IL-1β, IL-8, IL-6, and TNF-α circulating levels.

Then, we used a linear mixed effect model (LME) [6,7] with two fixed and two random effects (both slope and intercept) on the COVID-19 HD longitudinal data. For each individual and for each of the 10 selected features, the estimated fixed and random effects describe the feature progression on the overall cohort (the fixed effect) and the individual variation (the random effect). Supplementary figure 1 shows the individual LME fits.

Finally, to identify potential different clusters of COVID-19 HD patients, we performed a k-means clustering analysis [9] using as input the 20 features returned by the LME model (slope and intercept for each of the 10 features) and distinguish the different clusters, following a silhouette analysis with Dice distance.

**Statistical methods for the validation cohort**

For each subject of the validation cohort, we considered the baseline value of 8 potential “risk-factor” variables: sex, age, CRP, neutrophil, and lymphocyte percentages, procalcitonin, LDH, clinical presentation severity. As standard analysis in cluster validation, we used these variables to compute the Euclidean distances between each subject and the cluster centroids to assign each new patient to cluster 1 or 2.

**Supplementary tables**

**Supplementary Table 1**

**Full dataset of baseline clinical and laboratory parameters collected from COVID-19 positive and COVID-19 negative HD patients**

| **Clinical parameters** | **Laboratory parameters**  **(on blood)** | **Circulating cytokines levels** | **Cytometric analysis:**  **Profiles of CD4+ T helper (Th)** | **Cytometric analysis:**  **Maturation status of CD8+**  **and CD4+ T cells** | **Cytometric analysis:**  **Activation status of CD8+**  **and CD4+ T cells** |
| --- | --- | --- | --- | --- | --- |
| Age | Serum creatinine | IL-1β | Tfh/CD4+ | Naïve CD45RA+CCR7+CD95-/CD8+ | CD38+DR+/CD8+ |
| Sex | BUN | TNF-α | Tfh CCR4+/CD4+ | CD8+TSCM/CD8+ | CD38+DR+/CD4+ |
| Dialysis vintage | WBC | IL-6 | Tfh CXCR3+/CD4+ | CM CD8+:  CD45RA-CCR7+CD28+/CD8+ |  |
| Diabetes | Neutrophil count | IL-8 | Tfh CCR6+/CD4 | TM CD8+:  CD45RA-CCR7-CD28+/CD8+ |  |
| CVD disease | Lymphocyte count |  | CD161+CCR6+/CD4+ | EM CD8+:  CD45RA-CCR7-CD28-/CD8+ |  |
| COPD/asthma | CRP |  | Th1 CCR6-CCR4-CXCR3+/CD4+ | TE CD8+:  CD45RA+CCR7-CD28-/CD8+ |  |
| Clinical severity* | Procalcitonin, |  | Th2CCR6-CCR4+CXCR3-/CD4+ | Naïve CD45RA+CCR7+CD95-/CD4+ |  |
|  | Ferritin |  | Th17CCR6+CCR4+CXCR3-/CD4+ | CD4+TSCM/CD4+ |  |
|  | LDH |  | Th17CCR6+CCR4+CXCR3-CD161+/CD4+ | CM CD4+:  CD45RA-CCR7+CD28+/CD4+ |  |
|  | Albumin |  | Th1-Th17 CCR6+CCR4-CXCR3+/CD4+ | TM CD4+:  CD45RA-CCR7-CD28+/CD4+ |  |
|  | D-Dimer |  | Th1-Th17 CCR6+CCR4-CXCR3+CD161+/CD4+ | EM CD4+:  CD45RA-CCR7-CD28-/CD4+ |  |
|  |  |  | Th9CCR4-CCR6+/CD4+ | TE CD4+:  CD45RA+CCR7-CD28-/CD4+ |  |
|  |  |  | Th22CCR10+CCR4+CCR6+/CD4+ |  |  |

Abbreviations: hemodialysis, HD; cardiovascular disease, CVD; chronic obstructive pulmonary disease, COPD; white blood cell count, WBC; C-reactive protein, CRP; lactate dehydrogenase, LDH; interleukin, IL; Tumor necrosis factor-alfa, TNF-𝛼; cluster of differentiation, CD; central memory, CM; transitional memory, TM; effector memory, EM; terminal effector, TE; T-memory stem cells, TSCM.

* Clinical presentation severity was scored as reported in the text

**Supplementary Table 2**

**Comparisons between baseline characteristics of COVID-19 positive and COVID-19 negative HD patients**

|  | **COVID-19 positive HD** | **COVID-19 negative HD** | **p** |
| --- | --- | --- | --- |
| **N** | 16* | 6 |  |
| **Age, years** | 66.7 ±12.3 | 70.0±9.4 | 0.6 |
| **Sex, M/F** | 8/8 | 3/3 | 1 |
| **Dialysis vintage, months** | 51.5±46.0 | 46.3±26.5 | 0.6 |
| **Diabetes, N (%)** | 5 (31) | 3 (50) | 0.6 |
| **CVD disease, N (%)** | 8 (50) | 4 (66) | 0.6 |
| **COPD/asthma, N (%)** | 2 (12.5%) | 0 | 1 |
| **WBC, x10^9^/L** | 4.9±1.4 | 7.3±1.9 | 0.007 |
| **Lymphocytes, x10^9^/L** | 0.7±0.4 | 1.1±0.3 | 0.07 |
| **Neutrophils, x10^9^/L** | 3.7±1.4 | 5.6±1.7 | 0.03 |
| **Lymphocytes, (% WBC)** | 73.6±12.0 | 76.1±5.4 | 0.8 |
| **Neutrophils, (% WBC)** | 16.0±7.6 | 14.7±3.4 | 0.9 |
| **CRP, mg/L** | 39.8 (27.0-66.3) | 4.5 (3.0-11.7) | 0.006 |
| **Ferritin, µg/L** | 812 (591-1236) | 284 (227-408) | 0.02 |
| **D-dimer, μg/L** | 4731±11183 | 739±1330 | 0.04 |
| **Albumin, g/L** | 34±4 | 37±2 | 0.05 |
| **TNF-𝛂, pg/ml** | 48.0±16.0 | 22.9±4.0 | <0.001 |
| **IL-6, pg/ml** | 22.8 (15.1-40.2) | 5.2 (3.3-8.3) | 0.006 |
| **IL-8, pg/ml** | 52.5 (23.6-71.3) | 20.2 (9.0-32.0) | 0.02 |
| **IL-1b, pg/ml** | 1.1±0.9 | 0.4±0.0 | 0.009 |
| **ThfCCR4+/CD4+** | 0.3±0.2 | 1.9±1.4 | 0.04 |
| **Th2CCR6-CCR4+CXCR3-/CD4+** | 1.8±0.8 | 4.6±2.2 | 0.02 |
| **Th17CCR6+CCR4+CXCR3-/CD4+** | 1.7±1.1 | 3.6±0.9 | 0.003 |
| **Th9CCR4-CCR6+/CD4+** | 21.2±5.4 | 29.5±5.3 | 0.01 |
| **Th22CCR10+CCR4+CCR6+/CD4+** | 0.01±0.01 | 0.04±0.03 | 0.06 |
| **Th17CCR6+CCR4+CD161+/CD4+** | 1.4±1.0 | 2.8±1.0 | 0.02 |
| **CD38+DR+/CD8** | 16.0±9.1 | 6.9±4.6 | 0.02 |
| **CD8+TSCM/CD8+** | 1.7±1.5 | 0.7±0.5 | 0.04 |

Data are expressed by mean ± standard deviation (SD) or median-interquartile ranges (IQR) if they were not normally distributed.

* Baseline data were available in 16/17 COVID-19 HD patients enrolled in this study.

Abbreviations: hemodialysis, HD; cardiovascular disease, CVD; chronic obstructive pulmonary disease, COPD; white blood cell count, WBC; C-reactive protein, CRP; lactate dehydrogenase, LDH; interleukin, IL; Tumor necrosis factor-alfa, TNF-𝛼; T helper, Th; T-memory stem cells, TSCM.

**Supplementary Table 3**

**Baseline characteristics of the derivation cohort of COVID-19 positive HD patients**

**according to cluster assignment**

|  | **Cluster 1** | **Cluster 2** | **p** |
| --- | --- | --- | --- |
| **N** | 7 | 8* |  |
| **Age, years** | 68.1±15.6 | 64.4±10.7 | 0.7 |
| **Sex, M/F** | 1/6 | 7/1 | 0.01 |
| **Dialysis vintage, months** | 32.6± 19.6 | 72.8± 56.3 | 0.1 |
| **WBC, x10^9^/L** | 5.4± 1.3 | 4.4± 1.5 | 0.33 |
| **Lymphocytes, x10^9^/L** | 1.1±0.5 | 0.4±0.2 | 0.012 |
| **Neutrophils, x10^9^/L** | 3.8±1.4 | 3.5± 1.5 | 1 |
| **Lymphocytes, (% WBC)** | 21.0±7.5 | 11.8±4.9 | 0.03 |
| **Neutrophils, (% WBC)** | 66.6±13.8 | 78.6±6.2 | 0.06 |
| **CD8+TSCM/CD8+** | 0.6±0.4 | 2.3± 1.7 | 0.038 |
| **LDH, U/L** | 201.2± 42.3 | 323.0± 88.8 | 0.038 |
| **Procalcitonin, ng/ml** | 0.9± 0.5 | 3.3± 3.6 | 0.02 |
| **CRP, mg/L** | 22.2± 16.3 | 72.6± 44.1 | 0.017 |

* Baseline data were available in 8/9 COVID-19 HD patients included in Cluster 2

Abbreviations: hemodialysis, HD; white blood cells, WBC; C-reactive protein, CRP; lactate dehydrogenase, LDH; T-memory stem cells, TSCM.

**Supplementary Table 4**

**Validation cohort of longitudinal cluster analysis performed in COVID-19 positive HD patients: General characteristic and comparison between patients assigned to different clusters**

|  | **All patients** | **Cluster 1** | **Cluster 2** | | **p**  **Cluster 1 vs 2** |
| --- | --- | --- | --- | --- | --- |
| **N** | 30 | 16 | 14 | |  |
| **Age, years** | 73.3±16.3 | 75.7±15.5 | 70.4±17.3 | | 0.4 |
| **Sex, M/F** | 16/14 | 5 (29) | 3 (50) | | 0.4 |
| **Dialysis vintage, months** | 36 (14-71) | 43 (28-75) | 27 (7-64) | | 0.1 |
| **WBC, x10^9^/L** | 6.5± 4.9 | 5.4 ±5.1 | 7.7± 4.6 | | 0.02 |
| **Lymphocytes, x10^9^/L** | 0.7±0.3 | 0.9± 0.3 | 0.6±0.3 | | 0.03 |
| **Neutrophils, x10^9^/L** | 5.0±4.3 | 3.6±3.9 | 6.6± 4.4 | | 0.01 |
| **Lymphocytes, (% WBC)** | 15.4±9.5 | 21.2± 9.5 | 9.1±4.3 | | <0.0001 |
| **Neutrophils, (% WBC)** | 70.1±19.7 | 59.3±20.8 | 82.5± 6.9 | | <0.0001 |
| **LDH, U/L** | 234.0±72.1 | 207.9±49.1 | 260.1±83.4 | | 0.09 |
| **Procalcitonin, ng/ml** | 1.4 (0.6-4.6) | 0.8 (0.4-1.6) | 3.6 (1.2-9.5) | | 0.07 |
| **CRP, mg/L** | 14.7(4.0-39.0) | 8.0 (2.8-20.1) | 32.6 (12.1-50.1) | | 0.01 |
| **Clinical severity score >0, n (%)** | 26 (86) | 13 (80) | 14 (100) | | 0.04 |
| **Clinical severity score, n**   - **0** - **1**   **- 2** | 3  20  7 | 3  12  1 | 0  8  6 | | 0.018 |
| **Outcomes** |  |  |  | |  |
| **Death, n (%)** | 6 (20) | 1 (6.2) | 5 (35) | | 0.07 |
| **High-flow oxygen therapy, n (%)** | 4 (13) | 0 | 4 (28) | | 0.02 |
| **Duration of SARS-CoV-2 infection, days** | 21 (13-28) | 22 (14.5-35) | 20 (11-31) | | 0.38 |

Abbreviations: hemodialysis, HD; white blood cells, WBC; C-reactive protein, CRP; lactate dehydrogenase, LDH

**Supplementary Table 5**

**Reagents used for immunofluorescence analyses**

| **Type of marker** | **Specificity** | **Clone** | **Fluorochrome** |
| --- | --- | --- | --- |
| Cell lineage | CD3 | UCHT1 | BV786 |
|  | CD4 | RPA-T4 | APC-H7 |
|  | CD8 | RPA-T8 | PE-Cy7 |
| Maturation stage | CD45RA | H100 | BV605 |
|  | CD197(CCR7) | 150503 | PE-CF594 |
|  | CD95 | DX2 | BV711 |
|  | CD27 | M-T271 | BB515 |
| Th subsets | CXCR5 | RF8B2 | BV650 |
|  | CD196(CCR6) | 11A9 | BV421 |
|  | CCR10 | 1B5 | PerCP.Cy5,5 |
|  | CCR4 (CD194) | 1G1 | PE |
|  | CD183(CXCR3) | 1C6 | PE-Cy7 |
|  | CD161 | DX12 | APC |
| Activation | DR | G46-6 | PerCP.Cy5,5 |
|  | CD38 | HiT2 | PE |

Abbreviations: PE –phycoerythrin; Cy – cyanine; BV – brilliant violet; FITC – fluorescein; APC- Allophycocyanin; PerCP-Peridinin-chlorophyll proteins; BB-brilliant blue.

**Legend to supplementary Figure 1**

Individual linear mixed model fits. Each plot represents the individual LME fits estimated by the model for a specific feature of interest. In each plot, each line represents the fit of a specific COVID-19 HD subject, and colours are consistent among plots.

Abbreviations: white blood cell count, WBC; C-reactive protein, CRP; interleukin, IL; Tumor necrosis factor-alfa, TNF.

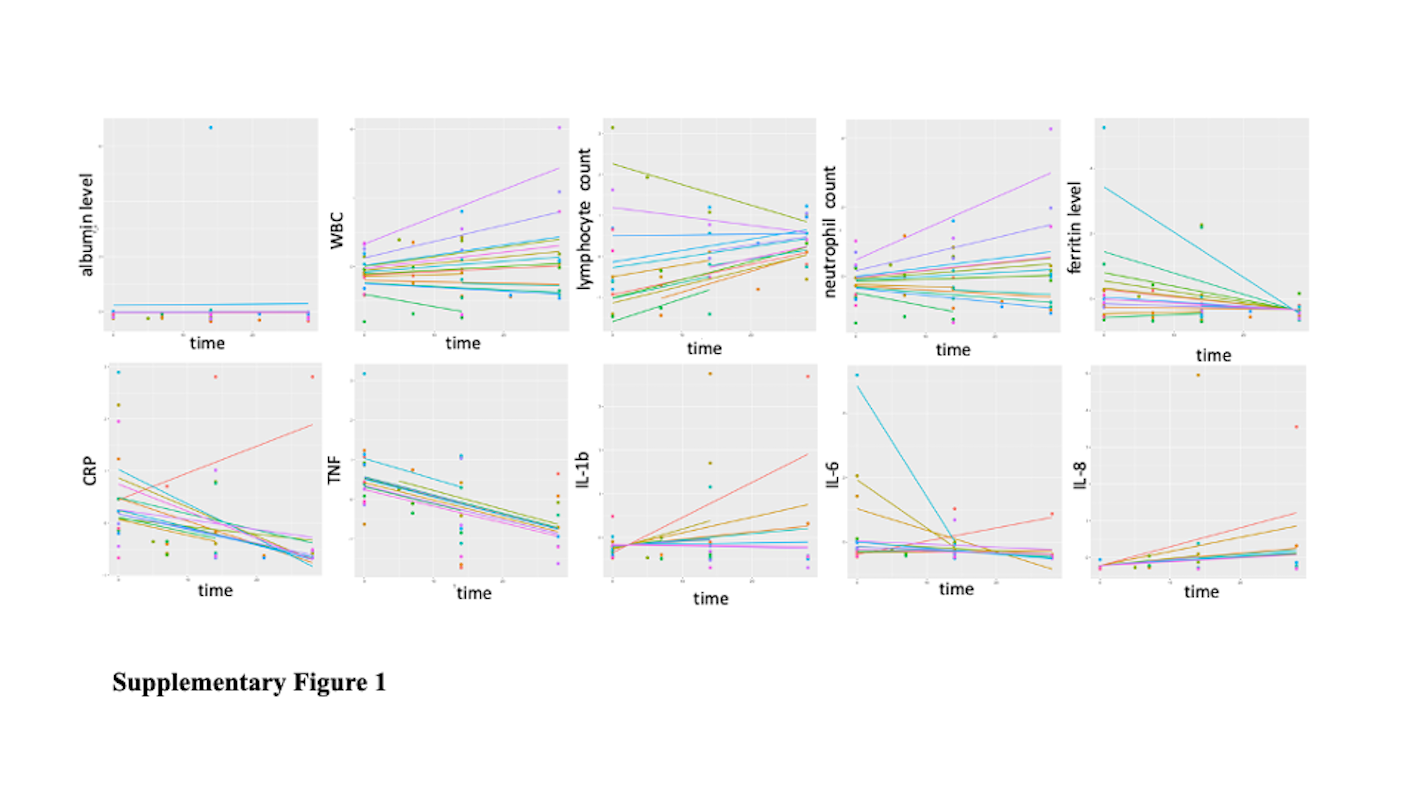
